# Prospective examination of mental health in university students during the COVID-19 pandemic

**DOI:** 10.1101/2021.07.29.21261196

**Authors:** Ru Jia, Holly Knight, Kieran Ayling, Carol Coupland, Jessica Corner, Chris Denning, Jonathan Ball, Kirsty Bolton, Joanne R Morling, Grazziela Figueredo, David Ed Morris, Patrick Tighe, Armando Villalon, Holly Blake, Kavita Vedhara

**Author notes:** Author to whom all correspondence should be addressed: Professor Kavita Vedhara, School of Medicine, University of Nottingham, University Park, Nottingham, NG7 2RD, UK. **Declarations**. **Funding** The study was funded by the Medical Research Council (Reference: MC_PC_20027, Principal Investigator J.B.) and the Institute for Policy and Engagement at the University of Nottingham (QR Duning Award, H.B., J.R.M., K.V.). JRM receives salary support from a Medical Research Council Clinician Scientist Fellowship [grant number MR/P008348/1]. The sponsors had no involvement in the study design, the collection, analysis and interpretation of data, or the preparation of the article. The views expressed are those of the authors and not necessarily those of the funders. **Ethics approval** Ethical approval was received from the University of Nottingham Faculty of Health Sciences Research Ethics Committee (FMHS 96-0920). **Consent to participate** Informed consent was obtained from all participants for being included in the study. **Consent for publication** Not applicable. **Availability of data and material** Data will be deposited in the University of Nottingham data archive. Access to this dataset will be embargoed for a period of 12 months in order for us to complete further analysis of the dataset. After that it may be shared with the consent of the Chief Investigator. **Code availability** Not applicable. **Author contributions** All authors designed the study. RJ analysed the data and RJ, HK and KV wrote the first draft. All authors provided critical revisions. All authors read and approved the submitted manuscript.

## Abstract

**Background:** The impact of changing social restrictions on the mental health of students during the COVID-19 pandemic warrants exploration.

**Aims:** To prospectively examine changes to university students’ mental health during the pandemic.

**Methods:** Students completed repeated online surveys at three time points (October 2020 (baseline), February 2021, March 2021) to explore relationships between demographic and psychological factors (loneliness and positive mood) and mental health outcomes (depression, anxiety, and stress).

**Results:** A total of 893 students participated. Depression and anxiety levels were higher at all timepoints than pre-pandemic normative data (*p*<.001). Scores on all mental health measures were highest in February, with depression and anxiety remaining significantly higher in March than baseline. Female students and those with previous mental health disorders were at greatest risk of poor mental health outcomes. Lower positive mood and greater loneliness at baseline were associated with greater depression and anxiety at follow-ups. Baseline positive mood predicted improvement of depression and anxiety at follow-ups.

**Conclusion:** Depression and anxiety were significantly higher than pre-pandemic norms, with female students and those with previous mental health difficulties being at greatest risk. Given these elevated rates, universities should ensure adequate support is available to meet potentially increased demand for services.

## Introduction

Young adults have been shown across many studies to have been at the greatest risk of mental health difficulties. (Jia, Ayling, Chalder, Massey, Broadbent, Morling, et al., 2020; Odriozola-González et al., 2020; Shanahan et al., 2020) During the Covid-19 pandemic, university students, as a subgroup of young people whose education, as well as other aspects of life, were disrupted were reporting increased levels of anxiety, depression, stress worldwide, during the initial phase of lockdowns. (Meda et al., 2021; Odriozola-González et al., 2020) The social restriction measures, however, changed frequently due to fluctuating levels of infections and mortality, serving as additional stressors. The Office for National Statistics (ONS) data suggested that in November 2020 more than half (57%) of the students reported slightly worsened wellbeing and mental health since starting the Autumn term, which increased to 63% in January. (ONS) Given the fluctuations in restrictions and their subsequent impact on students’ lives, it is therefore important to understand not only the prevalence of mental health difficulties during the various phases of the pandemic, but also the determinants of who is at greatest risk of mental health difficulties, as well as those who remain resilient. According to stress and coping theory, we would not expect effects on mental health to be uniform but to vary according to an individual’s cognitive appraisal and available psychosocial resources. (Folkman et al., 1991) Indeed, evidence from early in the pandemic has identified that female gender and prior mental health disorders were risk factors of anxiety, depression, and other mental health difficulties among both the general population (Xiong et al., 2020) and university students. (Elmer et al., 2020; Meda et al., 2021) However, these are demographic and clinical risk factors which cannot be modified. There are likely to be other psychological, potentially modifiable risk factors, that could also be important in understanding the risk of mental health difficulties in students. For example, perceived loneliness is known to have increased among students (Bu et al., 2020; Meda et al., 2021) and has been shown in previous, non-pandemic, research to be associated with an increased risk of mental health difficulties. (Leigh-Hunt et al., 2017)

Thus, we report here on the mental wellbeing in a UK university student cohort over 3 periods during university term 2020-2021, with a view to examining the persistence of mental health difficulties, and the demographic and psychological predictors of changes in anxiety and depression over time.

## Materials and Methods

### Recruitment

Ethical approval was received from the University of Nottingham Faculty of Health Sciences Research Ethics Committee (FMHS 96-0920). Participants were not compensated for their initial participation but were offered £10 per person for completing two follow-up surveys. Students enrolled at the University of Nottingham aged 18 or above were invited to participate via recruitment emails and through a social media campaign. Recruitment took place between October 5^th^ 2020 and November 1^st^ 2020.

### Procedures

Participants provided informed consent online before completing the online survey (JISC Online Surveys). The baseline survey was administered between 5/10/2020 – 1/11/2020 (Autumn term 2020-2021, pre second lockdown in England). All participants were invited by email to complete a second survey in February (1/2/2021-26/2/2021, Spring term 2020-2021, post-exam, 4 weeks into the third lockdown in England). Those who completed the second survey were invited to complete a final survey in March (10/3/2021 – 26/3/2021, Spring term 2020-2021, 9 weeks into the third lockdown in England).

At baseline, we collected data on sociodemographic characteristics (age, gender, stage of study, international status, pre-existing physical health issues, previous diagnosis of mental health disorders). In February and March we asked participants to report if they had to self-isolate at any point. At all time points we measured mental health outcomes including depression (Patient Health Questionnaire (PHQ-9, baseline α= 0.90), anxiety (Generalized Anxiety Disorder Scale (GAD-7, baseline α= 0.92), and stress (Perceived Stress Scale (PSS-4, baseline α= 0.70) and other psychological factors including positive mood (Scale of Positive and Negative Experience-Positive, SPANE-P, baseline α=0.93), loneliness, COVID-19 worry for themselves and close others. (Sheldon Cohen, 1988; Diener et al., 2010; Kroenke et al., 2010; Spitzer et al., 2006) Measures of sociodemographic characteristics, loneliness, positive mood, COVID-19 worry are described in Supplementary Appendix S1. Due to changes in social restriction measures before and after the university winter break, we expected some of the student to be unable to return to the university. Therefore location status was collected both in February and March in three categories: returned home for winter break and have not yet returned to the university, returned home for winter break but have returned to the university, never left the university during winter break.

### Statistical analysis

We first summarised the outcome variables (depression, anxiety and stress scores) and participant characteristics with appropriate descriptive statistics and examined histograms and scatterplots for normality. Depression and anxiety scores were square-root transformed due to non-normal distribution. Comparisons with pre-pandemic normative values were made using independent samples t-tests. (R. D. Kocalevent et al., 2013; B. Löwe et al., 2008) We then carried out analyses dichotomising depression and anxiety scores according to established thresholds for ‘caseness’ of depression and anxiety (PHQ-9 score greater or equal to 10, GAD-7 score greater or equal to 8). (National Collaborating Centre for Mental Health, 2019) Comparisons of mental health outcomes at three time-points were made using one-way repeated measures ANOVA. Cochran’s Q tests were conducted to examine the differences in the proportions of depression and anxiety cases over time.

Multivariable linear regression models were used to explore the independent relationships of demographic factors (age, gender, ethnicity, level of study, previous diagnosis of mental health disorders) and baseline psychological factors (perceived loneliness, perceived risk of COVID-19, positive mood, COVID-19 worry), and depression, anxiety and stress scores in February and March separately. Location and experiences of self-isolation by February or March were also controlled for in these models. Assumptions of linear regression (normality and homoscedasticity of residuals, linearity with continuous variables) and presence of outliers were assessed graphically. Incidence of depression or anxiety referred to individuals who were not classified as cases for depression or anxiety at baseline but became cases by February or March. We used logistic regression to estimate odds ratios (ORs) with 95% confidence intervals for associations with incidence and improvement of depression and anxiety cases by February or March using demographic and psychological factors at baseline, relative to no change of non-cases or cases. Improvement of depression or anxiety referred to individuals who were depression or anxiety cases at baseline but subsequently became non-cases by February or March. Demographic and psychological factors in the previous multivariable linear regression models were also controlled for in the logistic regression analysis.

Statistical analyses were performed using STATA (Version 16) and GraphPad Prism (Version 7).

## Results

### Cohort characteristics

In total, 897 students were recruited. Four participants were excluded due to being <18 years. Of these 60% (n=540) completed a follow-up survey in February and 53% (n=476) completed all three surveys. The final cohort (n=476) was predominately undergraduate students (n=420, 88%) with an average age of 20.8 years (SD = 3.7) and 71% (n=335) of female participants. Comparisons showed that participants who were lost to follow-up did not significantly differ from the final cohort on any of the sociodemographic and baseline psychological factors, with the exception for baseline anxiety (median (IQR), lost to follow-up: 3.00 (0.00-7.00) vs completers: 3.00 (1.00-8.00), t=2.04, *p*=.042).

### Mental health over time

Figure 1 shows the means of depression, anxiety, and stress among people who completed all 3 surveys over time. There were differences in means over time for depression (*F*(2, 474)=31.85, *p*<.001), anxiety (*F*(2, 474)=33.87, *p*<.001), and stress (*F*(2, 474)=3.70, *p*=.03). Depression and anxiety showed similar patterns: being highest in February compared with baseline (depression: F(1, 475)=57.01, *p*<.001, anxiety: F(1, 475)=67.73, *p*<.001) and March (depression: F(1, 475)=20.63, *p*<.001; anxiety: F(1, 475)=6.41, *p*=.012), while levels at baseline were significantly lower than in March (depression: F(1, 475)=7.56, *p*=.006; anxiety: F(1, 475)=31.53, *p*<.001). For perceived stress, levels were higher in February compared with baseline (F(1, 475)=6.32, *p*=.012). No significant differences were found between March and either February (F(1, 475)=19.36, *p*=.06) or baseline (F(1, 475)=0.72, *p=*.40). Mean values were higher than published population norms among young adults for depression and anxiety (all *p*<0.001) at all three time-points but not stress (baseline *p*=.13, February *p*=.96, March *p*=.31). (R. D. Kocalevent et al., 2013; B. Löwe et al., 2008)

**Figure 1.**
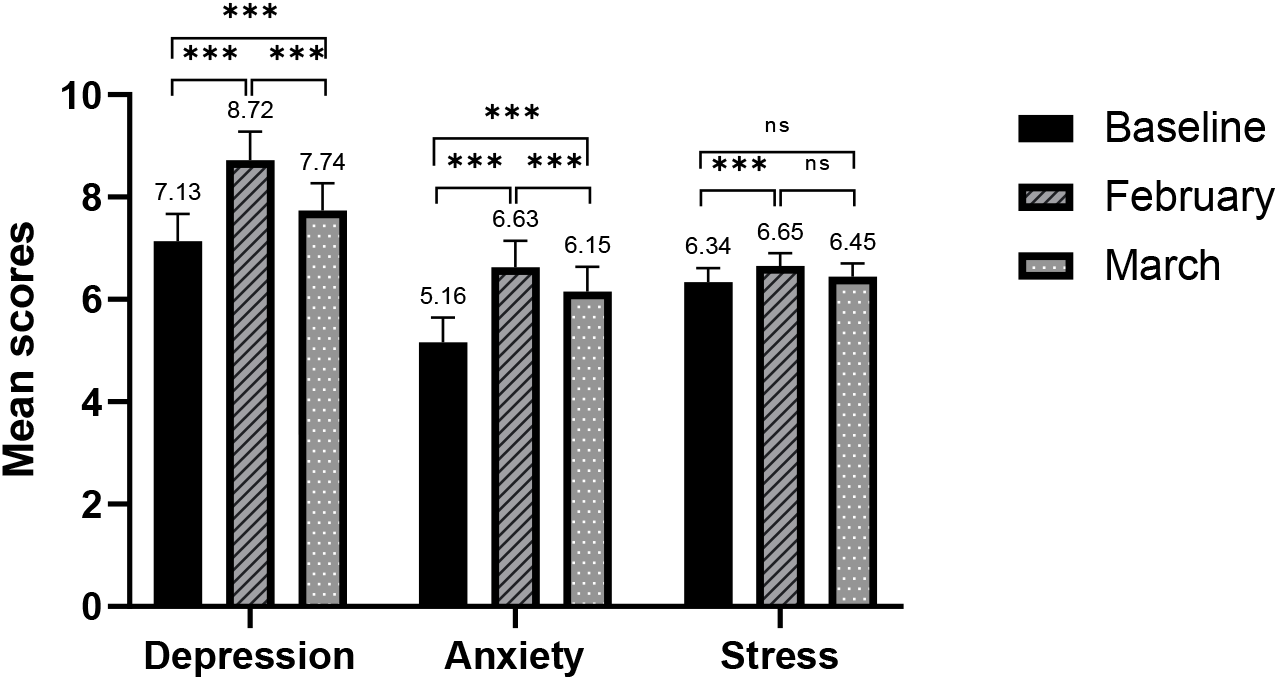
Mean scores for depression, anxiety, and stress over time among the final cohort (n=476). Bars are mean scores. Error bars are 95% confidence intervals. Score ranges: depression (0-27), anxiety (0-21), stress (0-16). Significant levels were based on repeated measure one-way ANOVA (square-root transformed mean scores for depression and anxiety, \*\*\**p*<.001).

When the scores were categorised into cases where thresholds for high-intensity psychological support in the NHS were met (i.e., score of 10 or greater for depression, and 8 or greater for anxiety), (National Collaborating Centre for Mental Health, 2019) results showed similar patterns. The prevalence of depression cases was lowest at baseline (29%), compared with February (37%, Cochran’s Q=18.29, *p*<.001) and March (33%, Cochran’s Q=4.05, *p*=.04), but no difference was observed between February and March (Cochran’s Q=3.31, *p*=.07). The prevalence of anxiety cases was lowest at baseline (22%), compared with in February (32%, Cochran’s Q=28.96, *p*<.001) and March (28%, Cochran’s Q=6.72, *p*=.01). The difference between February and March was also significant (Cochran’s Q=5.69, *p*=.02).

### Predictors of mental health outcomes

#### Positive mood, loneliness, and COVID-19 worry over time

Figure 2 shows the mean scores for positive mood, loneliness, and COVID-19 worry (for self and close others) at all three time-points. ANOVA analyses showed a statistically significant main effect for time on all psychological measures (positive mood: *F*(2, 474)=15.40, *p*<.001; loneliness: *F*(2, 474)=11.98, *p*<.001; COVID-19 worry for self: *F*(2, 474)=57.65, *p*<.001; COVID-19 worry for close others: *F*(2, 474)=48.44, *p*<.001). Post hoc testing revealed that mean scores for positive mood and loneliness in February were significantly different to baseline (positive mood: 20.11 vs 21.05, *p*<.001, loneliness: 2.15 vs 1.94, *p*<.001) and in March (positive mood: 20.11 vs 20.94, *p*<.001, loneliness: 2.15 vs 1.92, *p*<.001). However, the differences between baseline and March for both positive mood (21.05 vs 20.94, *p*=.58) and loneliness were non-significant (1.94 vs 1.92, *p*=.83). Mean scores for COVID-19 worry for both self (baseline: 1.97, February: 1.77, March: 1.62) and close others (baseline: 2.34, February: 2.21, March: 2.03) were significantly different between each two time-points (all *p*<.001).

**Figure 2.**
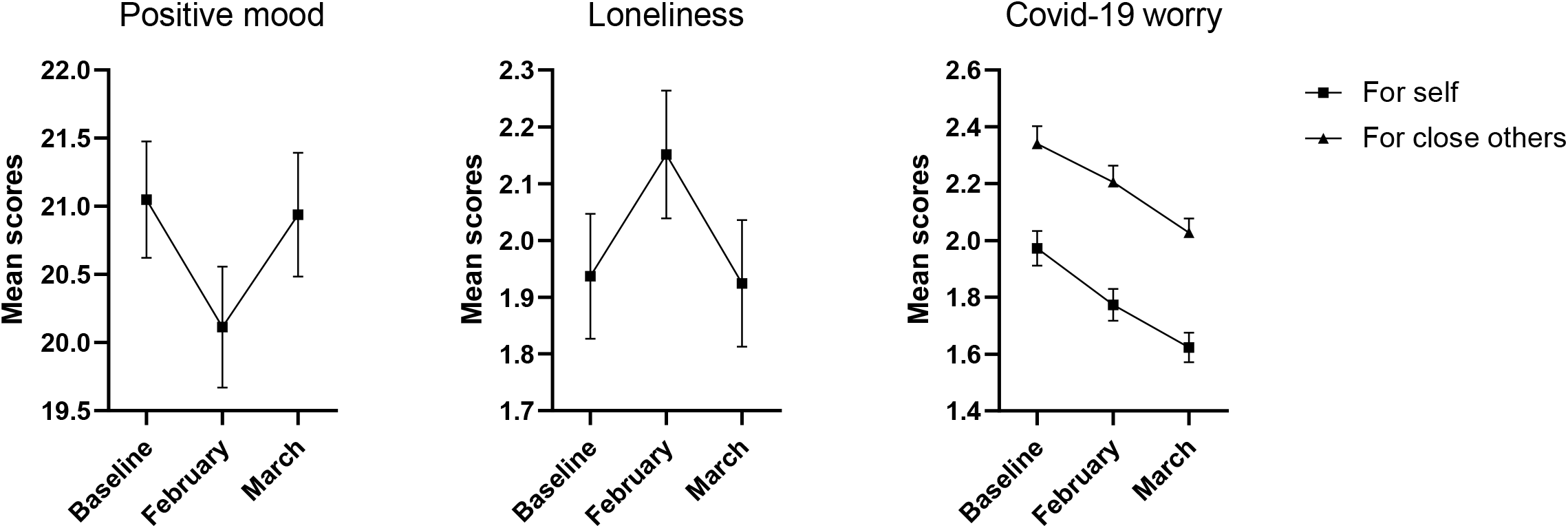
Mean scores for positive mood, loneliness, and COVID-19 worry (for self and others) over time among the final cohort (n=476). Bars are mean scores. Error bars are 95% confidence intervals. The mean scores for positive mood and loneliness in February were significantly different to baseline and March (p<0.001). The differences between mean scores at baseline and in March for positive mood and loneliness were non-significant (*p*>.05). Mean scores for COVID-19 worry for both self and close others were significantly different between each two time-points (*p*<.001).

#### Predicting spring term depression

Multivariable prospective regression analyses were conducted to explore sociodemographic and psychological predictors of mental health outcomes (Table 2). For depression, having previous diagnosis of a mental health disorder (February: B=0.43, CI: 0.22, 0.65; March: B=0.45, CI: 0.22, 0.69), lower positive mood at baseline (February: B=-0.10, CI: −0.12, −0.07; March: B=-0.09 CI: −0.12, −0.07) and greater loneliness at baseline (February: B=0.14, CI: 0.06, 0.22; March: B=0.15, CI: 0.06, 0.24) were significantly and independently associated with increased levels of depression in both February and March. Being female (B=0.29, CI: 0.09, 0.48) and staying at the university during the winter break (B=0.47, CI: 0.18, 0.75) were the only variables to be significantly associated with greater depression in February.

**Table 1:** Demographics among the final cohort and lost to follow-ups.

|  | Final cohort (n=476) | Lost to follow-up (n=417) |
| --- | --- | --- |
|  | N (%) | N (%) |
| **Age (n=892 Mean, SD)** | 20.8 (3.7) | 20.6 (3.0) |
| **Gender (n=889)** |  |  |
| Female | 335 (70.7%) | 221 (53.3%) |
| Male | 139 (29.3%) | 194 (46.7%) |
| **Ethnicity (n=893)** |  |  |
| White British | 334 (70.2%) | 256 (61.4%) |
| BAME background | 142 (29.8%) | 161 (38.6%) |
| **International students (n=893)** | 88 (18.5%) | 109 (26.1%) |
| **Year of study (n=893)** |  |  |
| 1<sup>st</sup> year Undergraduate | 116 (24.4%) | 117 (28.1%) |
| 2<sup>nd</sup> year Undergraduate | 95 (20.0%) | 72 (17.3%) |
| 3<sup>rd</sup> year Undergraduate | 138 (29.0%) | 122 (29.3%) |
| 4<sup>th</sup> Year Undergraduate | 60 (12.6%) | 46 (11.0%) |
| 5<sup>th</sup> year and above Undergraduate | 11 (2.3%) | 12 (2.9%) |
| Postgraduate | 55 (11.6%) | 39 (9.4%) |
| Other | 1 (0.2%) | 9 (2.2%) |
| **Existing physical health issue (n=888)** | 46 (9.7%) | 37 (8.2%) |
| **Previous diagnosis of mental health disorders (n=855)** | 114 (24.9%) | 85 (21.4%) |
| **Baseline depression score (n=893 Mean, SD)** | 7.1 (5.9) | 6.8 (6.2) |
| **Baseline anxiety score (n=893 Mean, SD)\*** | 5.2 (5.3) | 4.6 (5.2) |
| **Baseline stress score (n=893 Mean, SD)** | 6.3 (3.0) | 6.4 (2.9) |

\* Significant difference between completers and lost to follow-up

**Table 2:** Regression models showing prospective associations between sociodemographic and psychological explanatory variables at baseline and depression, anxiety and stress scores in February and March.

|  | Depression total score |  | Anxiety total score |  | Stress total score |  |
| --- | --- | --- | --- | --- | --- | --- |
|  | February | March | February | March | February | March |
|  | B (95% CI), *p* | B (95% CI), *p* | B (95% CI), *p* | B (95% CI), *p* | B (95% CI), *p* | B (95% CI), *p* |
| Age (per unit) | -0.0 (-0.05,0.00), 0.08 | -0.02 (-0.05, 0.01), 0.29 | -0.02 (-0.05, 0.01), 0.17 | -0.02 (-0.06, 0.01), 0.14 | -0.06 (-0.13, 0.01), 0.09 | **-0.08 (-0.16, -0.01), 0.03\*** |
| Female (Y/N) | **0.29 (0.09,0.48), 0.005\*\*** | 0.18 (-0.03, 0.39), 0.10 | **0.46 (0.25, 0.68), <0.001\*\*\*** | **0.29 (0.06, 0.51), 0.01\*** | **0.84 (0.34, 1.34), 0.001\*\*** | 0.36 (-0.17, 0.88), 0.18 |
| BAME background (Y/N) | 0.01 (-0.18,0.21), 0.89 | 0.03 (-0.18, 0.24), 0.78 | 0.07 (-0.14, 0.29), 0.50 | 0.11 (-0.11, 0.33), 0.33 | 0.31 (-0.20, 0.81), 0.23 | 0.63 (0.11, 1.15), 0.02* |
| Undergraduate (Y/N) | 0.02 (-0.28,0.33), 0.88 | 0.11 (-0.22, 0.44), 0.52 | 0.14 (-0.20, 0.48), 0.43 | -0.05 (-0.40, 0.29), 0.76 | 0.52 (-0.26, 1.29), 0.19 | -0.04 (-0.85, 0.78), 0.93 |
| Previous diagnosis of mental health disorder (Y/N) | **0.43 (0.22,0.65), <0.001\*\*\*** | **0.45 (0.22, 0.69), <0.001\*\*\*** | **0.49 (0.25, 0.73), <0.001\*\*\*** | **0.56 (0.32, 0.81), <0.001\*\*\*** | **1.09 (0.54, 1.64), <0.001\*\*\*** | **0.93 (0.35, 1.51), 0.002\*\*** |
| Positive mood (per unit) | **-0.10 (-0.12,0.07), <0.001\*\*\*** | **-0.09 (-0.12, -0.07), <0.001\*\*\*** | **-0.09 (-0.12, -0.07), <0.001\*\*\*** | **-0.09 (-0.11, -0.06), <0.001\*\*\*** | **-0.22 (-0.27, -0.17), <0.001\*\*\*** | **-0.22 (-0.28, -0.17), <0.001\*\*\*** |
| Perceived loneliness (per unit) | **0.14 (0.06,0.22), <0.001\*\*\*** | **0.15 (0.06, 0.24), <0.001\*\*\*** | **0.14 (0.05, 0.23), 0.003\*\*** | 0.10 (0.01, 0.19), 0.04 | 0.19 (-0.02, 0.40), 0.08 | 0.16 (-0.06, 0.38), 0.16 |
| COVID-19 worry for self (per unit) | 0.04 (-0.09,0.17), 0.52 | -0.02 (-0.16, 0.12), 0.82 | **0.21 (0.07, 0.36), 0.004\*\*** | 0.13 (-0.02, 0.28), 0.09 | **0.40 (0.07, 0.73), 0.02\*** | **0.42 (0.07, 0.76), 0.02\*** |
| Location<sup>b</sup> | February | March | February | March | February | March |
| Left university for winter break and have not yet came back | 0.11 (-0.08,0.30), 0.27 | 0.15 (-0.10, 0.40), 0.24 | -0.12 (-0.33, 0.09), 0.27 | 0.12 (-0.14, 0.38), 0.36 | 0.15 (-0.34, 0.64), 0.55 | 0.21 (-0.41, 0.83), 0.51 |
| Stayed at the university for winter break | **0.47 (0.18,0.75), 0.001\*\*** | 0.20 (-0.08, 0.48), 0.17 | 0.12 (-0.19, 0.44), 0.45 | 0.10 (-0.20, 0.39), 0.51 | **0.75 (0.03, 1.48), 0.04\*** | 0.62 (-0.08, 1.32), 0.08 |
| Self-isolation due to COVID-19 (Y/N) | 0.08 (-0.09,0.26), 0.36 | 0.04 (-0.14, 0.23), 0.65 | 0.02 (-0.17, 0.22), 0.83 | -0.01 (-0.21, 0.19), 0.95 | -0.06 (-0.51, 0.39), 0.80 | 0.23 (-0.24, 0.70), 0.33 |
|  | **Adjusted R<sup>2</sup>=0.36, n=456** | **Adjusted R<sup>2</sup>=0.30, n=456** | **Adjusted R<sup>2</sup>=0.34, n=456** | **Adjusted R<sup>2</sup>=0.28, n=456** | **Adjusted R<sup>2</sup>=0.31, n=456** | **Adjusted R<sup>2</sup>=0.26, n=456** |

\*\*\* $p < 0.001$ , \*\* $p < 0.01$ , \* $p < 0.05$
<sup>a</sup> A square-root transformation was applied to the dependent variable.
<sup>b</sup> Location at time of survey, comparison reference group “I left the university for winter break and have recently returned to the university”.

#### Predicting spring term anxiety

For anxiety, being female (February: B=0.46, CI: 0.25, 0.68; March: B=0.29, CI: 0.06, 0.51), having a previous diagnosis of mental health disorders (February: B=0.49, CI: 0.25, 0.73; March: B=0.56, CI: 0.32, 0.81), lower positive mood at baseline (February: B=-0.09, CI: − 0.12, −0.07; March: B=-0.09, CI: −0.11, −0.06) were significantly and independently associated with greater anxiety in both February and March. Greater loneliness at baseline (B=0.14, CI: 0.05, 0.23) and greater COVID-19 worry (B=0.21, CI: 0.07, 0.36) were significantly associated with greater anxiety in February.

#### Predicting spring term stress

For stress, having a previous diagnosis of mental health disorders (February: B=1.09, CI: .54, 1.64; March: B=0.93, CI: 0.0.35, 1.51), lower positive mood at baseline (February: B=-0.22, CI: −0.27, −0.17; March: B=-0.22 CI: −0.28, −0.17) and greater COVID-19 worry at baseline (February: B=0.40, CI: 0.07, 0.73; March: B=0.42, CI: 0.07, 0.76) were significantly and independently associated with greater stress in both February and March. Being female (B=0.84, CI: 0.34, 1.34) and staying at the university during the winter break (B=0.75, CI: 0.03, 1.48) were significantly associated with greater stress in February, while being younger (B=-0.08, CI: −0.16, −0.01) and from BAME background (B=0.63, CI: 0.11, 1.15) were significantly associated with greater stress in March.

#### Predictors of incident or improved depression and anxiety cases

We then explored risk factors predicting who became a case of depression or anxiety during the spring term, and those who became a non-case during this time (Table 3). Having previous diagnosis of mental health disorders (OR=2.45, 95% CI: 1.27, 4.70,), lower baseline positive mood (OR=0.90, 95% CI: 0.84, 0.97), and greater baseline loneliness (OR=1.32, 95% CI: 1.03, 1.69) were risk factors of incidence of depression cases, compared with those who were non-cases at all three time points. The results were slightly different for incidence of cases of anxiety: only female gender (OR=2.04, 95% CI: 1.14, 3.65) and lower baseline positive mood (OR=0.88, 95% CI: 0.83, 0.94) were independent risk factors. When looking at improved depression or anxiety cases, greater positive mood at baseline was a significant predictor for both depression and anxiety (depression: OR=1.11, 95% CI: 1.02, 1.22; anxiety: OR=1.15, 95% CI: 1.04, 1.28), compared with those who stayed cases at all time points. No previous diagnosis of mental health disorders was also a significant predictor of improved anxiety cases (OR=0.44, 95% CI: 0.20, 0.96).

**Table 3:** Logistic regression models showing associations between explanatory variables and incidence and prognosis of depression and anxiety cases^a^.

|  | Incident depression cases <sup>b</sup> | Incident anxiety cases <sup>b</sup> | Improved depression cases <sup>c</sup> | Improved anxiety cases <sup>c</sup> |
| --- | --- | --- | --- | --- |
|  | Odds Ratio (95% CI), *p* | Odds Ratio (95% CI), *p* | Odds Ratio (95% CI), *p* | Odds Ratio (95% CI), *p* |
| Age (per unit) | 0.97 (0.89, 1.05), 0.43 | 0.97 (0.90, 1.05), 0.48 | 0.96 (0.84, 1.08), 0.49 | 1.07 (0.92, 1.26), 0.37 |
| Female (Y/N) | 1.66 (0.94, 2.92), 0.08 | **2.04 (1.14, 3.65), 0.02\*** | 0.92 (0.40, 2.09), 0.84 | 0.59 (0.22, 1.63), 0.31 |
| BAME background (Y/N) | 0.86 (0.49, 1.51), 0.59 | 1.16 (0.67, 1.99), 0.60 | 0.83 (0.41, 1.69), 0.62 | 0.75 (0.33, 1.71), 0.50 |
| Undergraduate (Y/N) | 1.32 (0.52, 3.32), 0.56 | 0.81 (0.34, 1.92), 0.64 | 1.31 (0.41, 4.18), 0.64 | 1.16 (0.29, 4.62), 0.84 |
| Previous diagnosis of mental health disorder (Y/N) | **2.45 (1.27, 4.70), 0.007\*\*** | 1.65 (0.88, 3.11), 0.12 | 0.51 (0.25, 1.03), 0.06 | **0.44 (0.20, 0.96), 0.04\*** |
| Positive mood (per unit) | **0.90 (0.84, 0.97), 0.004\*\*** | **0.88 (0.83, 0.94), <0.001\*\*\*** | **1.11 (1.02, 1.22), 0.02\*** | **1.15 (1.04, 1.28), 0.007\*\*** |
| Perceived loneliness (per unit) | **1.32 (1.03, 1.69), 0.03\*** | 1.11 (0.87, 1.41), 0.40 | 0.83 (0.61, 1.14), 0.25 | 0.79 (0.55, 1.13), 0.71 |
| COVID-19 worry for self (per unit) | 1.07 (0.71, 1.62), 0.75 | 1.29 (0.86, 1.95), 0.22 | 0.92 (0.59, 1.42), 0.70 | 0.91 (0.56, 1.48), 0.42 |
| N | 338 | 365 | 164 | 133 |
| Pseudo R<sup>2</sup> | 0.11 | 0.09 | 0.10 | 0.13 |

\*\*\* $p < 0.001$ , \*\* $p < 0.01$ , \* $p < 0.05$
<sup>a</sup> A 'case' is defined as the PHQ-9 score greater or equal to 10 for depression, or the GAD-7 score greater or equal to 8 for anxiety, at which level someone would qualify for high-intensity psychological support in the National Health Service.
<sup>b</sup> Incident case refers to those who were not 'cases' at baseline and subsequently became a 'case' by February or March. The comparison groups were non-cases of depression and non-cases of anxiety at all 3 time periods, respectively.
<sup>c</sup> Improved case refers to those who were 'cases' at baseline and subsequently became 'non-cases' by February or March. The comparison groups were cases of depression and cases of anxiety at all 3 time periods, respectively.

## Discussion

We report here prospective findings from a longitudinal university student cohort established at the beginning of the 2020-2021 Academic Year in the midst of the COVID-19 pandemic. We observed that the overall levels of depression and anxiety significantly exceeded pre-pandemic normative data in similarly aged young adults at all time points. Of particular note was the fact that levels of depression, anxiety and stress all worsened in February 2021. It is possible that these increases in mental health difficulties were partly due to the strict and protracted lockdown measures since early January when most universities were delivering education virtually and students were unable to meet others outside their household. Despite the significant improvements by the end of March 2021, levels of depression and anxiety did not return to baseline levels. This suggests that the impact of the social restrictions on students’ mental health persisted throughout lockdown. However, it is also possible that students, like the general public, generally experience greater mental health difficulties in the winter when low mood and depressive symptoms are more prevalent. (Yang et al., 2010) Whether the increase in the mental health difficulties were mainly due to the lockdown restrictions or seasonal (Yang et al., 2010), our data showed concerning levels of mental health difficulties especially depression and anxiety in students. Universities should be aware of these difficulties and the likely increase in the demand for counselling services. Delivering adequate and timely psychological support to those in need is crucial. (Galea et al., 2020) Continued monitoring of students’ mental health may also be advantageous as students and the public traverse the next stages of the pandemic including the potential for a resumption of some restrictions in the winter months.

Another crucial element of dealing with the mental health difficulties is to identify who is at greater risk. In terms of demographic characteristics, we found that female gender was significantly associated with greater depression, anxiety and stress in February and anxiety in March. This echoes evidence from other cohorts in the pandemic and earlier evidence that seasonal depressive symptoms are more prevalent in women. (Elmer et al., 2020; Fancourt et al., 2021; Lyall et al., 2018) Having a previous diagnosis of mental health disorders was also a significant risk factor for depression, anxiety, and stress at follow-ups, which has also been identified as a risk factor in other cohorts. (Fancourt et al., 2021) Female students in our cohort also had double the odds of becoming an anxiety case, and having a previous diagnosis of a mental health disorder almost tripled the odds of becoming a depression case during follow-up, compared with those who remained non-cases. The consistency in these findings, both in our study and other cohorts (ONS) Jia, et al., 2020; ONS), suggests that individuals with these two characteristics are most vulnerable to future mental health difficulties. Universities are recommended to promote education or campaigns to raise awareness of mental well-being, especially in these groups. Interestingly, we also observed that students who stayed at the university during the winter break (or university closure period) were at increased risk of reporting greater depression and stress in February compared with those who went home and returned later. It is possible that these students were separated from their regular social interactions during and after the winter break partly due to lockdown measures, hence were lacking psychosocial resources to cope with potential mental health difficulties. This is also reflected by the increased loneliness and reduced positive mood in February in our results. It is important to note that, in a post-pandemic future, a proportion of students might still stay on campus during university closure periods. Although more evidence is needed, universities should be aware that these students might be at higher risks of mental health difficulties during this time therefore support should be made available especially in the winter months.

In terms of psychological risk factors, after taking demographic factors into account, we observed that lower positive mood at baseline was associated with depression, anxiety, and stress while greater baseline loneliness was associated with depression in both February and March, explaining 26-36% of the variance, in addition to the effect of other variables. These results were in line with other evidence at early stages of the pandemic. (Elmer et al., 2020; Jia, Ayling, Chalder, Massey, Broadbent, Coupland, et al., 2020; Meda et al., 2021) Students with lower positive mood or greater loneliness at baseline also had greater odds of developing depression in February and March, compared with those who were not cases at any timepoint. These results firstly echo risk factors identified by other studies (e.g., low social contact and emotional support) associated with negative mental health trajectories in students and the general population. (Elmer et al., 2020; Fancourt et al., 2021) Secondly, they suggest that loneliness and positive mood might mitigate the mental health sequela of COVID-19. Our analysis further showed that greater baseline positive mood was the only common predictor of improved cases of depression and anxiety during follow-up, compared with those who stayed cases at all time points. Taken together, it is suggested that positive mood might not only contribute to depression and anxiety, but might also act as a protective factor against deteriorations of mental health over time. (S. Cohen et al., 2003) Evidence has shown the effectiveness of positive psychology interventions (e.g., mindfulness and positive writing) in improving well-being and decreasing depressive symptoms among young people. (Sin & Lyubomirsky, 2009) Technology-based psychological interventions (e.g., digital mood-monitoring tools and virtual reality), as well as other interventions such as physical activity and animal interactions might also be effective in this group. (Barton & Pretty, 2010; Dubad et al., 2018; Grajfoner et al., 2017; Habak et al., 2020)

### Limitations

Despite the several strengths of this study such as longitudinal design and relatively large cohort, we acknowledge several limitations. First, the observational nature of the study means we cannot assume causal relationships between mental health outcomes and the impact of the COVID-19 pandemic. Second, nearly half (47%) of the original cohort were lost to follow-up. Although those who were lost to follow-up and completed all three surveys did not differ on most of the sociodemographic and baseline mental health measures, with the exception of anxiety. Although unlikely, it is possible that by excluding right censored data, our results overrepresented those with higher baseline anxiety. Third, our participants were self-selected and from a single institution in England with predominant proportion of undergraduate students and higher proportion of women than reported by Higher Education Policy Institute, (Higher Education Policy Institute) which also limits the generalisability of our findings. Nevertheless, our study showed the profound mental health difficulties university students experienced during the COVID-19 pandemic and point to potential avenues for psychological intervention (i.e., interventions addressing loneliness and positive mood).

## Data Availability

Data will be deposited in the University of Nottingham data archive. Access to this dataset will be embargoed for a period of 12 months in order for us to complete further analysis of the dataset. After that it may be shared with the consent of the Chief Investigator.

## Acknowledgements

We would like to thank all our participants as well as individuals and groups who provided help during our recruitment.

