## Supplementary Appendix for "Prospective examination of mental health in university students during the COVID-19 pandemic"

**Appendix S1: Details of measures**

|  | **Question/scale** | **Response(s)** |
| --- | --- | --- |
| **Sociodemographic charateristics** |  |  |
| *Age* | How old are you? | .. |
| *Gender** | What was your gender at birth? | Male |
|  |  | Female |
|  |  | Other |
|  |  | Prefer not to say |
| *Ethnicity** | What is your ethnicity | White – British, Irish, other |
|  |  | Asian/Asian British – Indian, Pakistani, Bangladeshi, other |
|  |  | Black/Black British – Caribbean, African, other |
|  |  | Chinese/Chinese British |
|  |  | Mixed race – White and Black/Black British |
|  |  | Middle Eastern/Middle Eastern British – Arab, Turkish, other |
|  |  | Mixed race – other |
|  |  | Other ethnic group |
|  |  | Prefer not to say |
| *Stage of study** | Which of the following best describe your current stage of study? | Foundation Course / Certificate |
|  |  | Registered for single modules only |
|  |  | 1^st^ year of undergraduate study |
|  |  | 2^nd^ year of undergraduate study |
|  |  | 3^rd^ year of undergraduate study |
|  |  | 4^th^ year of undergraduate study |
|  |  | 5^th^ year or higher of undergraduate study |
|  |  | I am a postgraduate |
| International status | Are you an international student (i.e., not from within the UK) | Yes, I’m from the EU |
|  |  | Yes, I’m from outside EU/UK |
|  |  | No, I’m from the UK |
| Previous diagnosis of mental health disorders | Do you have a history of anxiety, depression or any other mental health issue for which you have received treatment in the past? | Yes |
|  |  | No |
|  |  | Prefer not to say |
| *Pre-existing physical health issues* | Do you have an existing physical health issue that you think could affect your risk of getting COVID-19? | Yes |
|  |  | No |
|  |  | Prefer not to say |
| **Psychological factors** |  |  |
| *Perceived loneliness^†^* | How often have you felt lonely **over the past 2 weeks**? | Never (0) |
|  |  | Hardly ever (1) |
|  |  | Occasionally (2) |
|  |  | Some of the time (3) |
|  |  | Often/Always (4) |
| *COVID-19 worry (for self)* | Please read the following statements carefully and then select the one which best describe how you have felt over the past 2 weeks. | I do not worry about getting COVID-19. (1) |
|  |  | I occasionally worry about getting COVID-19.(2) |
|  |  | I spend much of my time worrying about getting COVID-19. (3) |
|  |  | I spend most of my time worrying about getting COVID-19. (4) |
| COVID-19 worry (for close others) | Please read the following statements carefully and then select the one which best describe how you have felt over the past 2 weeks. | I do not worry about my close relative(s)/ friend(s) getting COVID-19. (1) |
|  |  | I occasionally worry about my close relative(s)/ friend(s) getting COVID-19. (2) |
|  |  | I spend much of my time worrying about my close relative(s)/ friend(s) getting COVID-19. (3) |
|  |  | I spend most of my time worrying about my close relative(s)/ friend(s) getting COVID-19. (4) |
| *Positive mood^‡^* | Now we are going to show you some statements that people use to describe different feelings and emotions. Please read each statement and select the option that best reflects the way you felt over the past two weeks. | Very rarely or never (1)/ Rarely (2)/ Sometimes (3)/ Often (4)/ Very often or Always (5) |
|  | In the past 2 weeks, I have felt Positive. | Very rarely or never (1)/ Rarely (2)/ Sometimes (3)/ Often (4)/ Very often or Always (5) |
|  | In the past 2 weeks, I have felt Good. | Very rarely or never (1)/ Rarely (2)/ Sometimes (3)/ Often (4)/ Very often or Always (5) |
|  | In the past 2 weeks, I have felt Pleasant. | Very rarely or never (1)/ Rarely (2)/ Sometimes (3)/ Often (4)/ Very often or Always (5) |
|  | In the past 2 weeks, I have felt Happy. | Very rarely or never (1)/ Rarely (2)/ Sometimes (3)/ Often (4)/ Very often or Always (5) |
|  | In the past 2 weeks, I have felt Joyful. | Very rarely or never (1)/ Rarely (2)/ Sometimes (3)/ Often (4)/ Very often or Always (5) |
|  | In the past 2 weeks, I have felt Content. | Very rarely or never (1)/ Rarely (2)/ Sometimes (3)/ Often (4)/ Very often or Always (5) |

*Gender, ethnicity and stage of study were treated as binary variables in all analyses: gender (male, female), ethnicity (white British, non-white British), stage of study (undergraduate, not undergraduate).

^†^ The factors in *Italic* were hypothesised to be associated with an increased risk of adverse mental health outcomes and an increased risk of contracting COVID-19.

^‡^Positive mood was measured using the positive items from SPANE: Scale of Positive and Negative Experience (α=0.94).
